# Health-Related Quality of Life of Adults with Cutaneous Leishmaniasis at ALERT Hospital, Addis Ababa, Ethiopia

**DOI:** 10.1101/2023.02.26.23286484

**Authors:** Shimelis Doni, Kidist Yeneneh, Yohannes Hailemichael, Mikyas Gebremichael, Sophie Skarbek, Samuel Ayele, Abay Woday, Saba Lambert, Stephen L. Walker, Endalamaw Gadisa

**Affiliations:** ALERT Hospital, Addis Ababa, Ethiopia; Addis Ababa University, Addis Ababa, Ethiopia; Armauer Hansen Research Institute, NTD and Malaria Research Directorate, Addis Ababa, Ethiopia; Sheffield Teaching Hospitals NHS Foundation Trust, Sheffield, United Kingdom; Faculty of Infectious and Tropical Diseases, London School of Hygiene and Tropical Medicine, London, United Kingdom

## Abstract

**Background:** Cutaneous leishmaniasis (CL) is a growing public health threat in Ethiopia. *Leishmania aethiopica* is the predominant causative organism. Affected individuals develop chronic skin lesions on exposed parts of the body, mostly on the face, which are disfiguring and cause scarring. The effects of CL on the health-related quality of life (HRQoL) of affected individuals has not been formally assessed in Ethiopia.

**Objective:** To assess HRQoL in adults with active CL at ALERT Hospital, Addis Ababa, Ethiopia.

**Methods:** A cross-sectional study was done using the Amharic version of the Dermatology Life Quality Index (DLQI). Trained health staff administered the DLQI.

**Results:** Three hundred and two adults with active CL participated and all of them exhibited a reduced HRQoL. The median DLQI score was 10 (IQR 8). Almost half of the participants reported very poor HRQoL, 36.4% and 11.3% fell within the very large and extremely large effect categories respectively. DLQI scores were higher (median 18) in patients diagnosed with diffuse cutaneous leishmaniasis (DCL) compared to those with localized cutaneous leishmaniasis (LCL). The DLQI domain of ‘work and school’ was the most affected, scoring 73.3% and 66.6% of total possible core for female and male respectively, followed by that of ‘symptom and feeling’ (at 50.0% and 56.6% for female and male respectively). Men were more affected than women in the domains of ‘leisure’ (P=0.002) and ‘personal relationships’ (P=0.001). In the multivariate ordinal logistic regression site of lesion, clinical phenotype and age remained associated with significantly poor HRQoL.

**Conclusion:** The HRQoL impairment in people affected by CL is significant. This warrants improved care and treatment in Ethiopia, counseling on the nature of CL, therapeutic options as well as clinical outcomes and complications. Importantly, patient-reported outcome measures including the DLQI should be used to assess treatment efficacy patient care algorisms.

**Plain Language Summary:** Cutaneous leishmaniasis (CL) is an important public health problem in Ethiopia with an estimated incidence up to 50,000 cases per year. CL is predominately due to *Leishmania aethiopica. The transmission* is zoonotic by *Phlebotomus longipes* and *Ph. pedifer*, and hyraxes being the incriminated reservoir hosts. The lesion is chronic on the exposed part of the body, commonly on the face. Three main clinical phenotypes are recognized; localized (LCL), mucocutaneous (MCL), and diffuse CL (DCL) cutaneous leishmaniasis. The permanent damage and altered anatomy of the skin, nose, eyelids, ears and lips due to scarring is often associated with stigma.

This study aimed to assess the health-related quality of life (HRQoL) using the Amharic version of the Dermatology Life Quality Index (DLQI) in adults diagnosed with active CL. Our results show that the impact on HRQoL associated with CL is large. There was no significant difference between men and women, and urban and rural dwellers. Participants with DCL, the more extensive form of CL, had lower HRQoL compared to those with other forms. Those with lesions on their head and neck regions and younger than 50 year (20 to 49 years age group) had significantly low HRQoL (P<0.05).

The reduced HRQoL associated with CL in Ethiopia requires warrants improved, counseling on the nature of CL, therapeutic options as well as clinical outcomes and complications. Importantly, patient-reported outcome measures including the DLQI should be used to assess the efficacy of treatments and patient care algorisms.

## Introduction

The leishmaniases are neglected tropical diseases caused by an intracellular protozoan of the genus *Leishmania* transmitted by the bite of infected female phlebotomine sandflies. Cutaneous leishmaniasis (CL) is the most common form of leishmaniasis with an estimated million new cases per year globally [1]. CL, caused by *Leishmania aethiopica (L. aethiopica)*, is a major public health problem in Ethiopia with over 29 million people estimated to be at risk and up to 50000 new cases per year [2]. Affected individuals develop chronic, granulomatous cutaneous lesions on exposed parts of the body, predominantly the face. Lesions heal with scarring [3-5]. Children and young adults are the most affected [5-7]. The clinical phenotypes of CL due to *L. aethiopica* are localized (LCL), mucocutaneous (MCL) and diffuse (DCL) [8, 9]. MCL and DCL present significant therapeutic challenges.

CL has a severe psychosocial and economic impact, which contributes to perpetuating poverty. CL is associated with reduced health-related quality of life (HRQoL) of affected individuals due to the appearance of active skin lesions or the permanent scarring on exposed body sites [10, 11]. HRQoL encompasses physical activity, psychological well-being, degree of independence and social relationships, all of which are affected by skin disease [12]. Studies in other settings have shown affected individuals with CL have reduced quality of life. Boukthir et al., in a Tunisian study reported the impact of CL that ranged from scar related stigma to suicidal thoughts from fear of being handicaped and stress caused by close relatives [10]. Khatami et al.documented feelings of sadness, guilt, anxiety, stigma and exclusion through indepth interviews of 12 Iranian individuals with CL [11]. CL is associated with a negative impact on personal relationships and self-esteem resulting in anxiety and depression. Bennis et al., highlighted that CL could adversely affect life choices such as marriageability in endemic communities [13].

The Dermatology Life Quality Index (DLQI) has been used to assess the HRQoL of individuals with CL in Iran [14], Brazil [15] and the United Kingdom [16]. The proportion of individuals with a moderate or greater impact of their HRQoL ranged from 42.7-70%. There are no studies reporting data on HRQoL of individuals with CL in Ethiopia. We wished to assess the HRQoL using the DLQI in a referral hospital setting in Ethiopia.

## Methods Study setting

This work was done at the dermatology department of ALERT Hospital, Addis Ababa, Ethiopia *Study population and data collection*

A cross-sectional study was performed between December 2018 and December 2021. Individuals aged 18 years or older diagnosed with active CL (confirmed by microscopy and/or culture) who gave written informed consent were enrolled. The treating health care professionals classified participants as having LCL, MCL or DCL. Demographic and clinical data including location of lesions were recorded on a data collection form. The sites of skin lesions were categorised into regions as being on the head and neck or the torso and/or limbs or both regions. The head and neck skin lesions were further categorised into those affecting the face (not the lips or nose), lips or nose.

The validated Amharic version of the DLQI [17] was completed for each participant by a trained interviewer. The DLQI is a 10-item questionnaire addressing six aspects of life (domains);symptoms and feelings (Questions 1 and 2), daily activities (Questions 3 and 4), leisure (Questions 5 and 6), work and school (Question7), personal relationships (Questions 8 and 9), and treatment (Question 10). The scores to each of the 10 items add up giving a total score ranging from 0 to 30. Individuals rate the impact of their dermatological condition in the past week as “not at all, scored 0”, “a little, scored 1”, “a lot, scored 2”, “very much, scored 3”, “not relevant, scored 0” or Question 7, ‘prevented work or studying’ scored 3”. The HRQoL impact is interpreted using the total DLQI score as 0 – 1 no effect at all, 2 – 5 small effect, 6 – 10 moderate effect, 11 – 20 very large effect and 21 – 30 extremely large effect. The scores for each domain are expressed as a percentage of the total possible score for the domain. A higher DLQI scores means greater impairment on the patient’s health related quality of life (HRQoL).

### Data management and analysis

Data were checked for completeness and double entered into *EpiData* (version 3.1, *EpiData* Association, Odense, Denmark), exported and analyzed using *Stata* (version 17.0, *Stata* Corporation, College Station, TX, USA). A comparison of scores between two groups was made using the Mann–Whitney U inspected rank test, and for three or more groups the Kruskal–Wallis H test. Adjusted multivariable ordinal logistic regression analysis was performed to identify the independent predictors of effect of CL on HRQoL. P-values less than 0.05 were considered statistically significant.

### Ethical consideration

This study was reviewed and approved by AHRI/ALERT Ethics Review Committee (protocol number: PO/48/18). The participant information sheet and consent form were available in Amharic. Written informed consent was sought from all participants.

## Results

### Characteristics of the participants

Three hundred and two individuals diagnosed with active CL were recruited. The median age of participants was 29 years (IQR 21; 45), 56.0% male (169/302) and 66.6% urban dwellers (201/302). The body region most affected was the head and neck in 270 (89.4%), of which involve the nose in 151 (50%), other sites on the face in 122 (40.4%), and the lips in 35 (11.6%). The proportion of clinical phenotypes were 62.6% LCL (189/302), 34.4% MCL (104/302), and 3.6% DCL (9/302) (Table 1).

**Table 1:** Sociodemographic, disease characteristics and Dermatology Life Quality Index (DLQI) scores of participants

| Variables | Number (%) | DLQI Score |  |
| --- | --- | --- | --- |
|  |  | Median | IQR |
| Age (years) |  |  |  |
| 18-19 | 46 (15.2) | 9.5 | 10.0 |
| 20-29 | 106 (35.1) | 11 | 8.0 |
| 30-39 | 46 (15.2) | 14 | 9.0 |
| 40-49 | 42 (13.9) | 11 | 9.0 |
| ≥50 | 62 (20.5) | 8 | 5.0 |
| Sex |  |  |  |
| Male | 169 (56.0) | 11 | 9.0 |
| Female | 133 (44.0) | 10 | 8.0 |
| Residence |  |  |  |
| Urban | 201(66.6) | 10 | 8.0 |
| Rural | 101 (33.4) | 11 | 10.0 |
| Body region affected |  |  |  |
| Head and neck only | 270 (89.4) | 10 | 8.0 |
| Trunk and /or limbs only | 15 (5.0) | 8 | 6.0 |
| Multiple body regions | 17 (5.6) | 11 | 11.0 |
| Clinical phenotype |  |  |  |
| LCL | 189 (62.6) | 9 | 8.0 |
| MCL | 104 (34.4) | 11 | 9.5 |
| DCL | 9 (3.60) | 18 | 11.0 |

**Table 2.** Male and female participants with clinical phenotype of cutaneous leishmaniasis

| Sex of participants | Clinical phenotype (n, %) |  |  |
| --- | --- | --- | --- |
|  | LCL | MCL | DCL |
| Male | 104 (55.03) | 58 (55.77) | 7 (77.78) |
| Female | 85 (44.97) | 46 (44.23) | 2 (22.22) |
| Total | 189 (100) | 104 (100) | 9 (100) |

In terms of participants’ sex and clinical phenotype of CL, the males accounted for 55.5% (104/189), 55.77% (58/104) and 77.78% (7/9) of the LCL, MCL and DCL cases respectively (Table2).

### DLQI scores of participants

The overall median DLQI score for the study participants was 10 (IQR 8). The range of DLQI scores was 2 to 29. Median DLQI scores were higher in participants diagnosed with DCL (median 18) compared to participants with MCL (median 11) and LCL (median 9) (Table 1 and Figure 1). Similarly, participants in the 30-39 year age group had higher DLQI scores (median 14; IQR 9).

**Figure 1:**
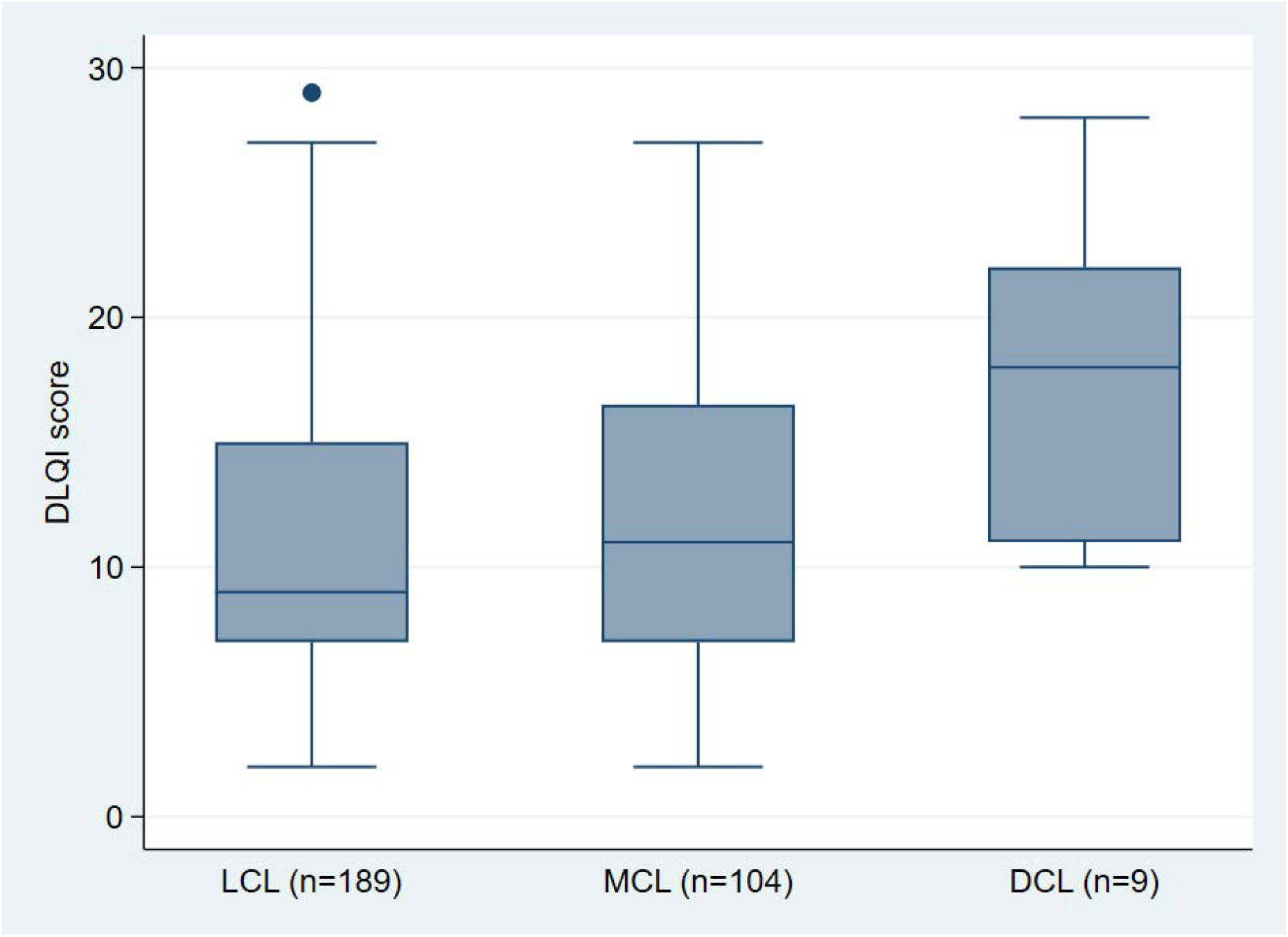
DLQI score associated with clinical phenotype of cutaneous leishmaniasis

### Size of HRQoL effect associated with active cutaneous leishmaniasis

The size of the HRQoL effect of active CL as measured by DLQI score ranged from small effect to extremely large effect (Table 3). Almost half of the participants reported very poor HRQoL, 36.4% and 11.3% fell within the very large and extremely large effect categories respectively. In addition, 39.4% of the participants experienced a moderate effect. Exceptionally, all the nine individuals with DCL had a DLQI score of 10 or more (Figure 1).

**Table 3:** The size of effect on HRQoL associated with clinical phenotype of cutaneous leishmaniasis.

| HRQoL effect | Clinical phenotype N (%) |  |  | Total N (%) |
| --- | --- | --- | --- | --- |
|  | LCL | MCL | DCL |  |
| Small | 25 (13.2) | 14 (13.5) | 0 (0) | 39 (12.9) |
| Moderate | 85 (45.0) | 33 (31.7) | 1 (11.1) | 119 (39.4) |
| Very large | 65 (34.4) | 41 (39.4) | 4 (44.4) | 110 (36.4) |
| Extremely large | 14 (7.4) | 16 (15.4) | 4 (44.4) | 34 (11.3) |

### DLQI domains

The percentage of domain scores were calculated from the total possible score. The percentage and the medians/ Interquartile ranges for each of the six domains is shown in Table 4. The ‘work and school’ domain had a high (73.3% and 66.6% for females and males respectively) total possible scores whereas the ‘personal relationships’ domain had low (16.6% and 25.0%, for females and males respectively). The HRQoL impairment was more pronounced in males compared to females in the ‘leisure’ (P=0.002) and ‘personal relationships’ (p= 0.001) DLQI domains.

**Table 4.**
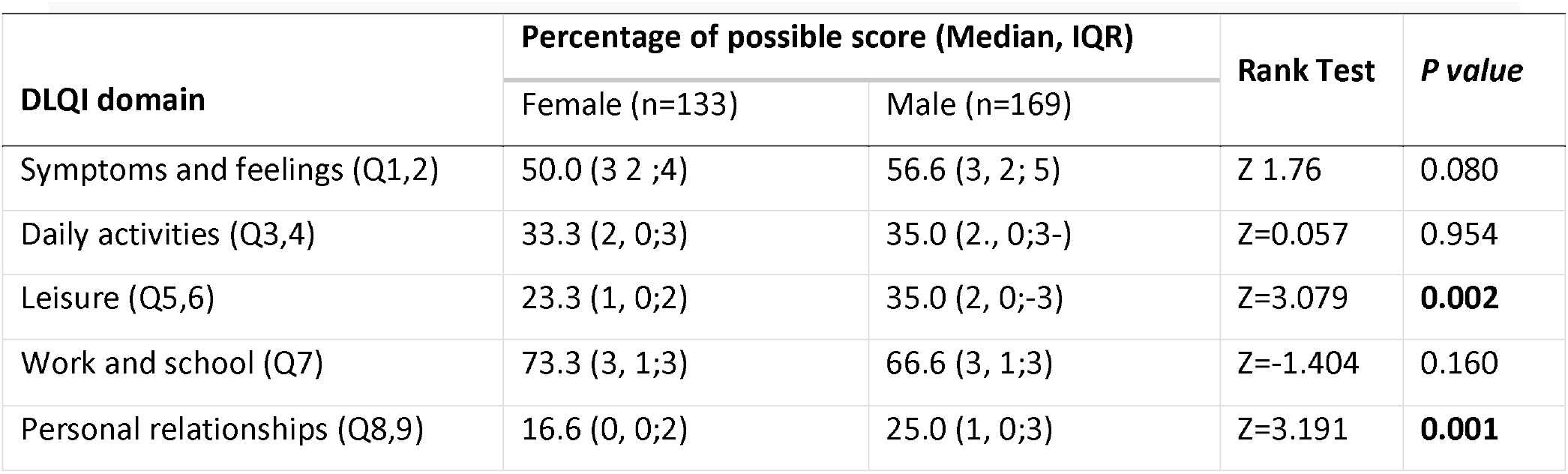
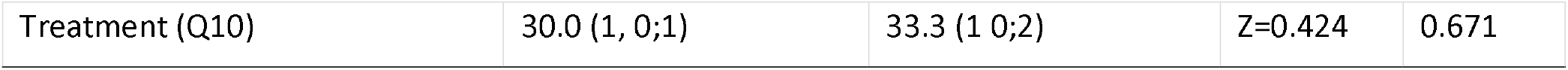
Percentage of total possible domain score and median/ Interquartile range of DLQI domains scores by sex of participants.

### Multivariate ordinal logistic regression

In the multivariate, clinical phenotype of CL, age of participants and the affected body region remain significantly associated with DLQI scores (Table 5). The odds of having low HRQoL was eight times higher in DCL cases (P=0.003) compared to those with LCL. Younger, 20 to 49 years, age groups and those having their head and face region affected had higher odds of having very poor HRQoL compared to those in 50 years and above, and those that have lesions on their Trunk and/or limbs. There was no significant difference in DLQI scores between male and female participants (P=0.260) or rural and urban dwellers (P=0.354).

**Table 5:** Multivariate ordinal logistic regression of DLQI score with sociodemographic and disease characteristic participants

| Variable | Median(IQR) | COR(95%CI) | AOR(95% CI) | P-Value (AOR) |
| --- | --- | --- | --- | --- |
| **Age in years** |  |  |  |  |
| 18-19 | 9.5(10) | 1.9(1.0,3.7) | 1.8(0.9,3.5) | 0.082 |
| 20-29 | 11(8) | 2.2(1.3,3.8) | 2.1(1.2,3.8) | **0.007** |
| 30-39 | 14(9) | 3.7(1.9,7.4) | 3.6(1.8,7.3) | **<0.001** |
| 40-49 | 11(9) | 2.7(1.4,5.3) | 2.7(1.3,5.3) | **0.004** |
| 50 and above | 8(5) | 1 | 1 |  |
| **Sex** |  |  |  |  |
| Male | 11(9) | 1.4(0.9,2.2) | 1.2(0.8,1.9) | 0.260 |
| Female | 10(8) | 1 | 1 |  |
| **Residence** |  |  |  |  |
| Urban | 10(8) | 0.7(0.5,1.1) | 0.8(0.5,1.2) | 0.354 |
| Rural | 11(10) | 1 | 1 |  |
| **Body site affected** |  |  |  |  |
| Multiple body parts | 11(11) | 4.0(1.2,13.6) | 2.2(0.6,7.7) | 0.195 |
| Head and neck | 10(8) | 2.0(0.8,4.9) | 3.2(1.2,8.4) | **0.018** |
| Trunk and/or limbs | 8(6) | 1 | 1 |  |
| **Clinical phenotype** |  |  |  |  |
| LCL | 9(8) | 1 | 1 |  |
| MCL | 11(9) | 1.4(0.9,2.1) | 1.3(0.8,2.0) | 0.162 |
| DCL | 18(11) | 5.7(1.7,18.4) | 8.0(2.0,31.5) | **0.003** |

## Discussion

Skin diseases exert a considerable impact on social relationships, psychological status and on the daily routine of patients. Cutaneous leishmaniasis, caused mainly by *Leishmania aethiopica*, is (re)emerging public health challenge in Ethiopia. The disease causes severe disfigurement, because of either destruction of and/or formation of lifelong scars on aesthetically important body parts [8].

The concept of HRQoL encompasses physical activity, psychological well-being, the patient’s degree of independence and his/her social relationships. Various studies showed the impact of CL in HRQoL of affected individuals [14, 15, 18]. In this article, we presented the effect of CL on patients’ HRQoL, as measured by the DLQI, at ALERT Hospital, in Ethiopia.

In this study the first of its kind involving large number of participants, the median DLQI score was 10 (IQR 8). All participants with active CL experienced an effect on their HRQoL, which ranged from small effect to extremely large effect. Almost half of the participants reported very poor HRQoL, 36.4% and 11.3% fell within the very large. This is higher HRQoL impact compared to studies conducted in other part of the world. A study done in Belo Horizonte, Brazil in 2009-2010 showed that among 20 CL patients 70 % of them had a moderate/ large impact on their HRQoL [14]. Another study done in UK on returning travelers indicated that 63% among 46 adults reported moderate or higher impact on their quality of life [15]. The high proportion of our participants (270, 89.4%) having lesion in their head and neck regions, aesthetically important region, and the significantly poor HRQoL (P=0.018) observed in them might explain the large proportion of patients with high HRQoL effect.

The younger age group (20-49) had greater odds of having poor HRQoL compared to the 50 or above age group a finding that trend with a finding in Psoriasis patients [19]. Probably, for younger age group visible aesthetic damages might more crucial while attempting to enter or assume more social, or interpersonal relationship.

Despite the differences in sociocultural, study setting and causative species difference people are concerned with the level of aesthetic damage due to CL. The CL clinical phenotype were associated significantly with different DLQI scores. Individuals with DCL had 8.8 times higher odds having poor HRQoL compared to patients with LCL. Probably, patients feel the level of disfigurement. In line to our finding Refai et al., observed that in active LCL cases those with Plaque-ulcers type had higher DLQI score [20]. This is also consistent with studies conducted in Iran by Vares et al., which indicated that those with ulcerated lesions had lower quality of life compared to non-ulcerated [14].

We witnessed that the “Leisure” (P=0.002) and “Personal Relationship” (P=0.001) DLQI sub domain significantly correlated with male sex. This observation corroborates the previous explanations that the culturally assumed social status of males might probably made them more concerned of their HRQoL. Moreover, in a population based cohort study on schizophrenia, it was observed that Ethiopian women did not reported upfront on their overall life satisfaction or quality of spousal relationships [21].

The high CL related HRQoL impact in the “symptom and sign”, and “work and School” DLQI sub domain and severe form of CL, DCL, trends with previous studies [14, 15]. The highest impact of CL in Vares et al.,[14] study was in the “symptom and sign” sub domain while it was in the “work and School” in that of Toledo et al., [15]. Treatment appears to have a low effect perhaps the study participants were treatment naïve and were yet to experience the effect of treatment on their HRQoL.

This study showed that active CL holds important HRQoL burden on affected individuals. Thus, warrants improved, counseling on the nature of CL, therapeutic options as well as clinical outcomes and complications related to CL care and management. Importantly, patient-reported outcome measures including the DLQI should be used to assess the efficacy of treatments and patient care algorisms.

### Limitations of the study

This study is a cross sectional prior to treatment, we cannot comment on possible treatment related changes in HRQoL. The clinical classification was based on the judgement of the dermatologist who assessed the patient rather than standardized case definitions.

## Data Availability

Available upon request

## Acknowledgements

We are grateful to the study participants for their time and the dermatologists and staff of the Dermatology Department. Dr Michael Marks participated in discussions concerning analysis. A special thank you to Edom Getachew who assisted with data entry.

